# Accurate Stride Length Prediction from Proximal IMU Sensors Using a Compact Linear Model

**DOI:** 10.64898/2026.01.26.26344854

**Authors:** Ren Gibbons, Jennifer Yee, Rebecca Webster, Doug Wajda

## Abstract

**Objective:** Accurate stride length measurement is essential for assessing functional mobility, yet gold-standard methods remain confined to laboratory settings. This study aimed to develop and validate a computationally efficient, interpretable linear model for predicting stride length using thigh- and shank-mounted inertial measurement units integrated into a wearable neuromodulation sleeve.

**Methods:** Data from the sleeve were collected from 29 healthy adults performing walking bouts at four self-selected speeds. Participants traversed a pressure-sensitive gait mat, providing gold standard labels. A linear regression model was developed from engineered features from the kinematics data streams and validated against a held-out test set (*n* = 6) using leaveone-participant-out cross-validation.

**Results:** The final linear model utilized five predictors: participant height, shank range of motion (ROM), thigh ROM, and thigh swing duration metrics. It achieved high predictive accuracy with a mean absolute error (MAE) of 5.98 cm, a mean absolute percentage error (MAPE) of 4.53%, and an *R*^2^ of 0.89. The model significantly outperformed naive baseline models (*p <* 0.05) and performed similarly to more complex non-linear architectures, such as neural networks and random forests. Notably, 88.4% of strides were predicted within 10% of the ground truth.

**Conclusion:** A parsimonious linear model leveraging proximal limb kinematics provides accurate and biomechanically interpretable stride length estimation. Low computational demand makes it suitable for real-time, on-device gait monitoring in wearable assistive technologies, facilitating clinical assessments in real-world environments.

## 1. Introduction

Stride length is a widely used clinical and research measure of gait performance, reflecting functional mobility [1], disability, and mortality [2, 3]. Reliable measurement of stride length is important for both clinical monitoring and biomechanical research. With the growing popularity of wearable inertial sensors for gait assessments [4], quantitative gait assessment in naturalistic, non-laboratory environments is increasingly accessible [5]. However, the accuracy and replicability of wearable sensor–based stride length estimation remains limited by differences in algorithmic methods, sensor placement, and signal quality [6].

Gait mats remain the gold standard for stride length measurement but are restricted to the laboratory environment [7]. Optoelectronic motion capture also provides accurate measurement from worn sensors due to its high precision and reliability in capturing three-dimensional kinematics [8]. Yet, these systems require specialized equipment, controlled environments, and trained personnel [9, 10], making them costly and impractical for routine or longitudinal use. This has driven interest in wearable inertial measurement units (IMUs) as a portable and cost-effective alternative for measuring gait in real-world settings [4, 11].

A range of IMU-based algorithms have been proposed for stride length estimation. Traditional approaches such as zero-velocity update have been widely used [12, 13], but they rely on strict assumptions about foot-ground contact that often fail in non-normative gait and are prone to drift from double integration errors. Studies using machine learning algorithms have reported mean absolute percentage error (MAPE) as low as 1.7% averaged over hundreds of strides using a neural network [14] to approximately 5% [15] for single-sensor approaches, while multi-sensor configurations have achieved comparable accuracy [16]. However, many of these methods rely on computationally complex algorithms, which are not well suited for real-time, wearable use. Moreover, the diversity of sensor locations and processing pipelines complicates comparisons and limits generalizability across systems [6].

The objective of this study was to develop and validate a stride length prediction model using thigh-and shank-mounted IMU data. Although most gait analysis studies have used foot- or lower-back–mounted IMUs [11], sensor location remains an open design choice rather than a settled standard. Limb placements on the thigh or shank offer both biomechanical and practical advantages for stride length estimation. Biomechanically, a thigh-mounted IMU has been shown to outperform foot-mounted IMUs in hemiparetic populations [17]. Trunk-mounted sensors only indirectly capture the motion of the legs, which are the mechanical generators of stride. Practically, proximal sensors are less exposed to impact, less influenced by footwear or placement variability, and easier to don for users with impaired mobility. This study was conducted in a healthy population as foundational research, and future work will investigate suitability in populations with neurologically impaired gait.

Unlike many previous IMU-based stride length approaches that rely on distal sensors or computationally intensive nonlinear models, this study demonstrates that a small, biomechanically interpretable linear model using proximal limb kinematics can achieve comparable accuracy while remaining suitable for real-time embedded deployment.

## 2. Methods

### 2.1. Participants

Following Institutional Review Board approval (Cleveland State University, IRB-FY2024-253) and written informed consent in accordance with institutional guidelines and the Declaration of Helsinki, 29 healthy adults (15F/14M, age 28.8 ± 8.4 years, Table 1) were recruited via campus advertisements. Participants were required to be 18–64 years old and capable of 30 minutes of ambulation during a two hour period. Exclusion criteria included lower motor neuron disease, sensory loss in the legs, implanted cardiac devices, local malignancy or thrombosis, orthopedic injuries, or any history of impaired gait.

**Table 1.** Summary of participant demographics and clinical characteristics for the full cohort (*n* = 29) and the stratified training (*n* = 23) and test (*n* = 6) sets.

| Variable | All Participants<br>($n = 29$) | Training Set<br>($n = 23$) | Test Set<br>($n = 6$) |
| --- | --- | --- | --- |
| Sex (F/M) | 15 / 14 | 12 / 11 | 3 / 3 |
| Age (years) | $28.9 \pm 8.4$ | $27.6 \pm 6.8$ | $33.8 \pm 12.3$ |
| Height (cm) | $171.9 \pm 10.4$ | $173.0 \pm 10.5$ | $167.5 \pm 9.7$ |
| Weight (kg) | $72.1 \pm 14.1$ | $73.0 \pm 14.7$ | $68.6 \pm 12.3$ |
| Leg Length (cm) | $93.3 \pm 7.2$ | $94.8 \pm 6.9$ | $87.5 \pm 5.4$ |
| Dominant Leg (R/L) | 27 / 2 | 21 / 2 | 6 / 0 |

### 2.2. Apparatus

Each participant wore a Cionic Neural Sleeve on their self-reported dominant leg. The Neural Sleeve is an FDA-cleared Class II neuromodulation device designed to assist walking in individuals with upper motor neuron-related mobility impairments. A Control Unit, which houses the computing module and battery, connects via a short cable at the upper thigh and communicates with a smartphone application using Bluetooth Low Energy. Two IMUs are integrated on the lateral thigh and shank, streaming calibrated Euler angles.

Walking data were recorded using a Protokinetics Zeno 20-ft long by 4- ft wide pressure-sensitive gait mat placed on a level indoor surface with no obstructions.

### 2.3. Protocol

Participants donned the Neural Sleeve, and a standardized IMU calibration was conducted. Participants completed two 2.5-minute walking bouts at each of four self-selected speed conditions: Intermittent-Slow, Slow, Normal, and Fast. For the Intermittent-Slow condition, participants were instructed to walk as if “moving through an art gallery”, resulting in interspersed pauses, reflecting sub-normal walking speeds of clinical populations. For Slow, participants walked at a slow, steady pace. The Slow condition was collected only for the last 12 participants. Instructions for Normal were to walk at a normal, comfortable pace, and Fast instructions were “as quickly but safely as possible”. The gait mat was placed in the center of a 10-meter course, ensuring steady state gait on the mat. At the end of each pass, participants turned and walked back over the mat until completing the 2.5-minute bout. The order of conditions was randomized, and participants were offered up to a 5-minute seated rest between trials.

### 2.4. Data Acquisition

Full resolution IMU kinematic data from the Neural Sleeve were streamed at 100 Hz, logged to the Control Unit, and later used in offline stride-level analysis. IMU data include limb Euler angles computed from on-device calibrated quaternions. Corresponding spatiotemporal gait data from the Zeno mat were exported using the Protokinetics software suite and uploaded to a secure server.

### 2.5. Data Processing

Ground-truth stride lengths were extracted directly from the Zeno mat output files. The stride length prediction pipeline began with segmentation of the IMU kinematic data into individual strides. Segmentation events were computed using a peak-finding algorithm on the sagittal-plane component of the thigh Euler angle stream. Strides were segmented using a real-time, envelope-based peak-finding algorithm on the sagittal thigh Euler angles, which triggered on positive-to-negative derivative transitions exceeding a preset amplitude threshold. The algorithm also enforces a minimum waiting period between peaks.

IMU segmentation events were temporally aligned with gait mat heel-strike timestamps. Alignment was performed by systematically testing all possible temporal alignments between the two timestamp series. For each candidate offset, nearest-neighbor matches were identified under a monotonicity constraint to preserve event order. The offset yielding the smallest cumulative absolute timing error across matched pairs was selected. The resulting mean absolute synchronization error was 13.8 ± 10.7 ms.

Next, stride-level features were extracted from thigh and shank IMU segments using Python post hoc processing that emulates the embedded firmware environment. To ensure deployability on the microcontroller, computationally expensive operations such as median or standard deviation were intentionally excluded.

Stride-level features were derived from the sagittal Euler angle stream of the thigh and shank IMUs. For the thigh sensor, fourteen features were: peak-based duration metrics (peak-to-trough time, trough-to-peak time, peak-to-peak time - and their squared terms), amplitude measures (start stride peak, end stride peak, stride trough, start stride peak - stride trough, end stride peak - stride trough), and statistical summaries (stride sum, stride sample count, stride mean). “Start” and “end” refer to the peak that commences or completes a segmented stride. For the shank sensor, six complementary features were computed from the sagittal Euler angle stream: amplitude measures (stride max, stride min, stride max - stride min) and statistical summaries (stride sum, stride sample count, stride mean). Two anthropometric features (participant height and leg length) were also included; these were included to account for inter-individual biomechanical scaling effects known to influence stride length [18].

### 2.6. Model Development

Model development began with forward feature selection, in which the model architecture is an empty set of predictors and iteratively adds the single best-performing remaining feature. Performance was evaluated by mean absolute error (MAE). The smallest model that achieved near-optimal performance was selected to balance predictive accuracy and computational efficiency.

A range of regression and machine learning architectures were evaluated, including ordinary least squares (OLS) linear regression, ridge regression, lasso regression, random forest, gradient boosting, and a multilayer perceptron (MLP) neural network.

Model performance was assessed using a leave-one-participant-out cross-validation strategy. In each fold, data from one participant were withheld for testing while models were trained on data from the remaining participants. This ensured that the model’s performance generalized across individuals and was not biased by participant-specific gait characteristics.

Following extensive testing, a linear regression model with a small subset of features was selected as the final architecture. Despite its simplicity, this model achieved comparable or superior accuracy to more complex non-linear methods while requiring minimal memory and computational resources. The model is suitable for on-device deployment within the Neural Sleeve’s micro-controller and ensures interpretability of features, a key advantage for clinical and biomechanical applications.

### 2.7. Validation and Evaluation

A held-out test set of six participants (*n* = 6) was reserved for final model evaluation. These participants were randomly selected prior to model development and excluded from all training, cross-validation, and feature selection procedures. Their data were not examined until the model architecture was finalized. The test cohort included three men and three women, and participants were of a comparable age and height distribution to the full study sample (mean ± SD: 33.8 ± 12.3 years, 1.68 ± 0.1 m).

Model performance was evaluated using clinically interpretable error metrics defined a priori: MAE, MAPE, root mean squared error (RMSE), coefficient of determination (*R*^2^), and the percentage of strides within 5% and 10% of ground-truth stride length. Error thresholds of 5% and 10% were utilized to evaluate the proportion of “high-fidelity” predictions, aligning with the reported minimal detectable change for stride length in healthy cohorts [19]. These metrics were computed across all strides within the test set to assess absolute and relative accuracy.

To contextualize performance, four naive baseline models were included:

(1) linear regression on height only, (2) linear regression on stride time only, (3) linear regression on height and stride time, and (4) a simple mean predictor equal to the mean stride length of the training data. These baselines were selected to represent simple but physiologically plausible predictors.

Statistical comparisons between the final model to the other candidates and naive baselines were conducted using the Wilcoxon signed-rank test on participant-level MAE values. A nonparametric test was selected because the small sample size (*n* = 6) precluded reliable assessment of normality required for parametric tests such as the paired t-test.

## 3 Results

The final linear model achieved a test set MAE of 5.98 cm (MAPE = 4.53%) equal to or outperforming all other models.

The final linear model (Equation 1) included five predictors: height, shank max - min (i.e. shank ROM), thigh peak-to-trough time, thigh peak-to-trough time squared, and start thigh peak - thigh trough (i.e., thigh ROM). The final model scaler values and coefficients are shown in Table 2.

**Table 2.** Final model feature coefficients and descriptive statistics for predictors including participant height and inertial sensor-derived kinematic variables. P-values determine the significance each predictor.

| Feature | Mean<br>(units) | St Dev<br>(units) | Model Coef<br>$\beta_n$ | p-value |
| --- | --- | --- | --- | --- |
| Intercept (cm) | N/A | N/A | 137.98 | < **0.001\*** |
| Shank ROM (deg) | 77.99 | 6.88 | 8.69 | < **0.001\*** |
| Height (cm) | 172.38 | 10.26 | 11.11 | < **0.001\*** |
| Thigh P-T Time (sec) | 0.81 | 0.20 | -14.83 | < **0.001\*** |
| Thigh P-T Squared (sec<sup>2</sup>) | 1.00 | 1.31 | 4.86 | < **0.001\*** |
| Thigh ROM (deg) | 44.58 | 5.98 | 3.76 | < **0.001\*** |

Statistical significance was set at $\alpha = 0.05$ ; significant p-values are indicated in bold and marked with an asterisk (\*); St Dev = standard deviation; P-T Time = time from peak to trough of sagittal thigh Euler angle; ROM = range of motion

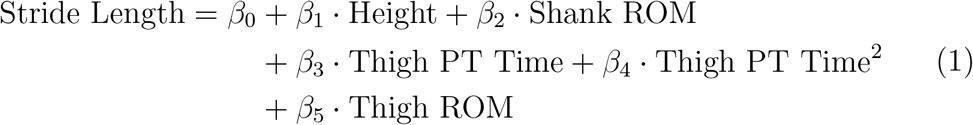

Height and shank ROM had the strongest positive influence on stride length, followed by the quadratic term for thigh peak-to-trough time and thigh ROM. Conversely, the linear thigh peak-to-trough term contributed negatively, indicating that longer swing durations are associated with shorter strides when other factors are held constant. Together, these relationships align with established gait mechanics, namely taller individuals and larger lower-limb excursions tend to exhibit longer strides [20].

Per stride results given in the Bland-Altman plot in Figure 1 are colored by participants. Intra-participant clusters reflect the various walking instructed speeds. While a small prediction bias of 2.85 cm exists, little proportional bias is observed, i.e. the bias is consistent across the range of stride lengths. Furthermore, the bias is skewed by Participant 3 who exhibits the largest over-prediction error.

**Figure 1.**
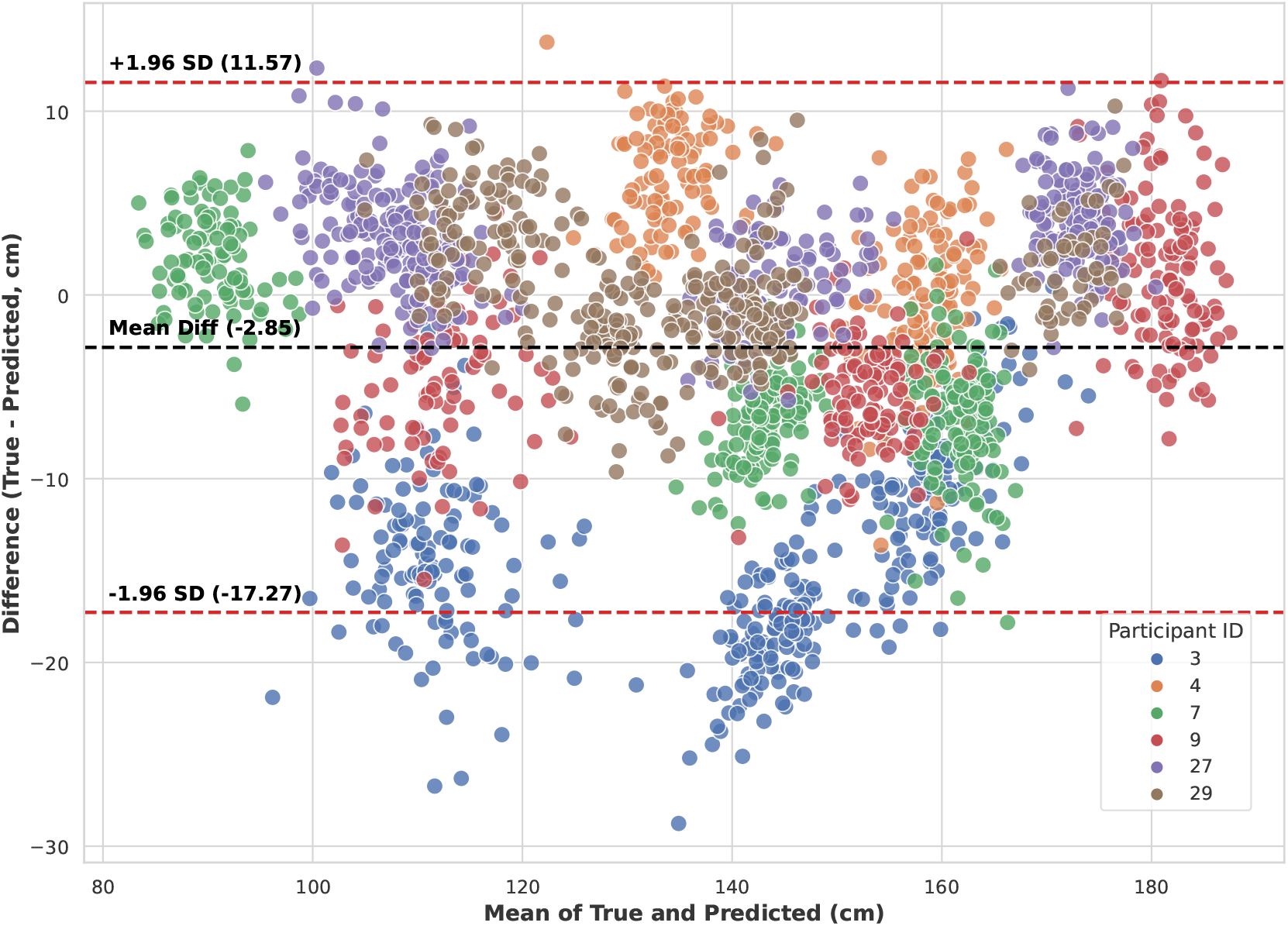
Bland-Altman plot comparing ground truth and predicted stride lengths for the test set across six participants. The solid black dashed line represents a small overestimation bias of 2.85 cm, though bias is consistent across the range of true stride lengths.

Across all architectures evaluated, OLS linear regression achieved the best overall performance. Regularized linear models (Ridge and Lasso) showed nearly identical results suggesting minimal feature collinearity. The model yielded MAE of 5.98 cm, MAPE of 4.53%, and RMSE of 7.89 cm. Additionally, 69.2% of strides were predicted within 5% error and 88.4% within 10% error of the ground truth. In contrast, more complex non-linear models such as random forest, gradient boosting, and MLP neural network offered little performance advantage, with MAEs ranging from 6.2-8.1 cm. The results are shown in Table 3.

**Table 3.** Comparison of predictive performance between the final linear regression model, alternative machine learning architectures (“Other Candidate”), and naive baseline models (“Base Case”) for stride length estimation.

| Model Type | Model | $R^2$ | MAE<br>(cm) | MAPE<br>(%) | RMSE<br>(cm) | Pct < 5%<br>MAPE | Pct < 10%<br>MAPE |
| --- | --- | --- | --- | --- | --- | --- | --- |
| Final | Final OLS | 0.90 | 5.98 | 4.53 | 7.89 | 69.19 | 88.41 |
| Other Candidate | Ridge | 0.90 | 5.97 | 4.52 | 7.89 | 69.24 | 88.41 |
| Other Candidate | Lasso | 0.90 | 5.98 | 4.51 | 7.96 | 72.46 | 88.61 |
| Other Candidate | MLP | 0.88 | 6.74 | 5.13 | 8.60 | 60.13 | 84.50 |
| Other Candidate | Gradient Boosting | 0.88 | 7.01 | 5.49 | 8.71 | 56.02 | 83.61 |
| Other Candidate | Random Forest | 0.88 | 7.02 | 5.37 | 8.75 | 56.12 | 88.46 |
| Other Candidate | Decision Tree | 0.84 | 7.96 | 6.08 | 9.90 | 49.33 | 84.00 |
| Other Candidate | Elastic Net | 0.84 | 8.11 | 6.38 | 10.12 | 48.74 | 81.03 |
| Base Case | Height +<br>Stride Time OLS | 0.77 | 9.26 | 6.95 | 11.97 | 46.81 | 75.48 |
| Base Case | Stride Time OLS | 0.65 | 12.32 | 9.49 | 14.76 | 31.30 | 61.61 |
| Base Case | Mean Stride | -0.00 | 20.63 | 15.86 | 25.12 | 23.87 | 39.08 |
| Base Case | Height OLS | 0.00 | 21.17 | 16.01 | 25.09 | 18.77 | 39.08 |

$R^2$ = coefficient of determination; MAE = mean absolute error; MAPE = mean absolute percentage error; RMSE = root mean square error; Pct < 5% MAPE and Pct < 10% MAPE represent the percentage of predictions with error below those respective thresholds; OLS = ordinary least squares regression; MLP = multilayer perceptron

The linear model similarly achieved superior predictive performance compared to all naive baseline models across all evaluation metrics. The best-performing baseline (linear regression on height and stride peak-to-peak time) achieved an MAE of 9.26 cm and only 46.8% of strides within 5% error. Simpler baselines, such as height-only and stride-time-only regressions, performed worse, with MAEs ranging from 12.3-21.2 cm and less than 31.3% of strides within 5% error.

Figure 2 shows MAE (cm) of the final OLS compared to MLP (best other candidate), and height+stride time OLS (best base case). Linear regression and MLP offer similar performance across speeds, and outperform the naive baseline for non-normal walking speeds.

**Figure 2.**
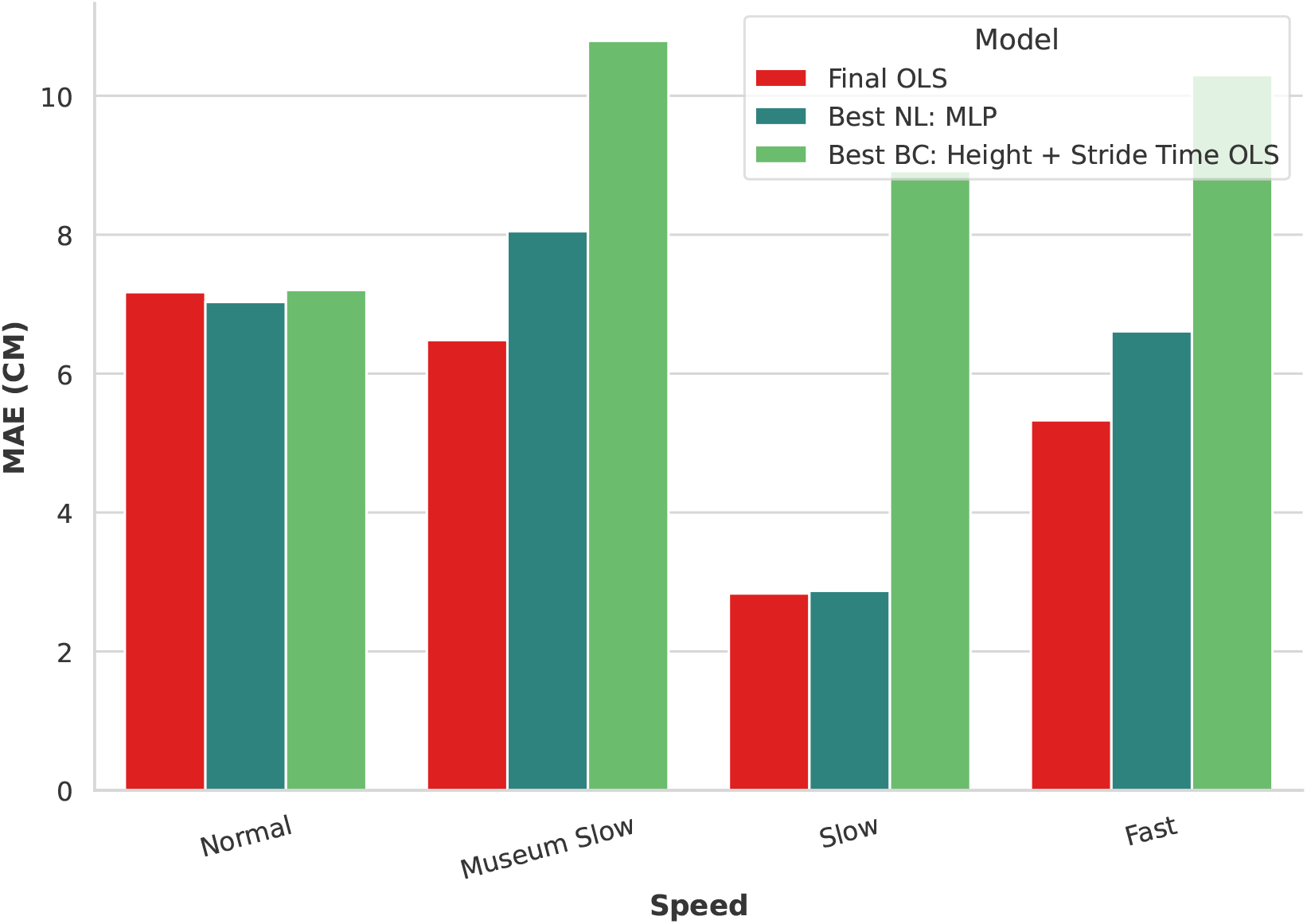
Comparison of MAE across different walking speeds for the Final Linear Regression (OLS) model, the best-performing non-linear model (MLP), and the best-performing baseline model (Height + Stride Time OLS). The Final OLS model demonstrates consistent predictive accuracy across all speeds, notably outperforming the baseline during all non-Normal speeds.

To confirm the statistical significance of these model performance differences, Wilcoxon signed-rank tests were performed comparing each candidate and baseline model to the final model. The final regression significantly out-performed all baseline comparisons at *α* = 0.05, as well as elastic net, decision tree, and random forest as shown in Table 4. These results indicate that the final linear model, though parsimonious, captured subject-independent structure in the data and generalized robustly across the test set of participants.

**Table 4.** One-sided Wilcoxon signed-rank test results comparing the final linear regression model against candidate and baseline models. Median mean absolute error (MAE) differences and corresponding inter-quartile range (IQR) are reported for the test set (*n* = 6 participants). P-values (*α* = 0.05) determine if comparison models exhibit significantly higher error than the Final OLS model.

| Model Type | Comparison | Median MAE<br>Diff (cm) | IQR MAE<br>Diff (cm) | Wilcoxon | p-value |
| --- | --- | --- | --- | --- | --- |
| Naive Baseline | Height +<br>Stride Time OLS | 2.87 | 1.60 | 21.0 | **0.0156\*** |
| Naive Baseline | Stride Time OLS | 6.98 | 7.38 | 21.0 | **0.0156\*** |
| Naive Baseline | Height OLS | 13.08 | 5.62 | 21.0 | **0.0156\*** |
| Naive Baseline | Mean Stride | 13.99 | 4.20 | 21.0 | **0.0156\*** |
| Other Candidate | Decision Tree | 2.21 | 0.95 | 20.0 | **0.0312\*** |
| Other Candidate | Elastic Net | 2.63 | 2.46 | 19.0 | **0.0469\*** |
| Other Candidate | MLP | 1.15 | 0.93 | 19.0 | **0.0469\*** |
| Other Candidate | Random Forest | 0.86 | 1.30 | 17.0 | 0.1094 |
| Other Candidate | Gradient Boosting | 0.50 | 0.98 | 17.0 | 0.1094 |
| Other Candidate | Lasso | 0.00 | 0.82 | 9.0 | 0.6562 |
| Other Candidate | Ridge | -0.00 | 0.00 | 3.0 | 0.9531 |

Statistical significance was set at $\alpha = 0.05$ ; significant p-values are indicated in bold and marked with an asterisk (\*); MAE = mean absolute error, IQR = interquartile range, OLS = ordinary least squares

## 4. Discussion

This study developed and validated a compact, interpretable linear model for estimating stride length using thigh- and shank-mounted IMUs in the Cionic Neural Sleeve. The model achieved high accuracy (MAE = 5.98 cm; MAPE = 4.53%), within the range of performance for published IMU algorithms [21, 22], while enabling on-device computation.

The model demonstrates that proximal limb kinematics sufficiently predict stride length. Height, thigh and shank ROM, and stride timing emerged as most predictive, aligning with established biomechanical relationships [22]. These features generalized robustly across participants. Further, nonlinear architectures (random forests, neural networks) did not improve accuracy, underscoring that well-chosen features can outperform model complexity.

Clinically, this approach enables real-time stride length estimation on wearable hardware, supporting continuous gait monitoring and adaptive control without cloud computation. The interpretable features provide insight into model behavior, while the thigh- and shank-based configuration reduces distal sensor burden, facilitating long-term wear.

Although the model generalized to unseen healthy participants, this work does not claim generalization to pathological or variable-terrain gait. Stride kinematics in populations with neurologically or orthopedically impaired gait may differ substantially. The present results should be interpreted as a foundational validation of the modeling approach, providing a basis for subsequent adaptation in clinical cohorts. Future work will explicitly investigate model evaluation and adaptation to support clinical research.

## 5. Conclusion

This study validated a compact linear model for estimating stride length from thigh- and shank-mounted IMUs in the Cionic Neural Sleeve. The model achieved accuracy comparable to nonlinear architectures while maintaining interpretability and real-time computational efficiency [21]. Predictive features aligned with known gait mechanics, reinforcing biomechanical validity and clinical transparency [22].

These findings demonstrate accurate stride length estimation using physiologically meaningful features from proximal limb sensors, without heavy processing or foot-mounted instrumentation [23]. This supports transparent, on-device gait analytics in wearable assistive technologies, enabling scalable gait monitoring and rehabilitation in real-world settings. Future work will validate the model in clinical populations with pathological gait to test generalizability and clinical utility.

## Declarations

### CRediT Authorship Contribution Statement

**Ren Gibbons**: Conceptualization, Formal analysis, Methodology, Software, Visualization, Writing - original draft. **Jennifer Yee**: Conceptualization, Methodology, Writing - review & editing. **Rebecca Webster**: Conceptualization, Methodology, Project administration, Writing - review & editing. **Doug Wajda**: Conceptualization, Data curation, Funding acquisition, Investigation, Methodology, Project administration, Writing - review & editing.

### Ethics Approval and Consent to Participate

This study was approved by the Cleveland State University Institutional Review Board (IRB-FY2024-253) and conducted in accordance with the Declaration of Helsinki. All participants provided written informed consent prior to participation.

### Conflict of Interest Statement

Cionic, Inc. is the manufacturer of the device used in this study, where Ren Gibbons, Jennifer Yee, and Rebecca Webster are employees. The Cionic equipment used for data collection was provided to Cleveland State University at no cost. Cionic, Inc. employees were involved in study design, predictive algorithm development, and writing of the manuscript as part of their regular employment duties. Cleveland State University received no financial compensation from Cionic, Inc. for the data collection associated with this study. All authors affirm that the study was conducted objectively and that all results are reported transparently.

### Funding Sources

This work was supported by Cionic, Inc. The sponsor contributed to the study design, data analysis, model development, and manuscript preparation.

### Data Availability Statement

Data will be made available upon reasonable request.

*Acknowledgments*

The authors thank all study participants for volunteering their time. We gratefully acknowledge the support of the Cleveland State University Health Sciences and Human Performance staff for assistance with data collection and logistics.

### Declaration of Generative AI Assistance in the Manuscript Preparation Process

During the preparation of this work, the author(s) used ChatGPT and Gemini to assist in the literature review and the initial drafting process. Following the use of these tools, the author(s) reviewed, edited, and verified the content as needed. All citations were manually checked for accuracy. The author(s) take full responsibility for the content of the published article.

## Notes

### Clinical Trial

NCT06503354

### Author Declarations

Institutional Review Board or Cleveland State University gave ethical approval for this work (IRB-FY2024-253)

